# Observer-rated outcomes of Communication-Centered Treatment (CCT) for adults who stutter: A preliminary study

**DOI:** 10.1101/2023.06.19.23290720

**Authors:** Courtney T. Byrd, Geoffrey A. Coalson, Danielle Werle

## Abstract

Previous studies have reported that adults who stutter demonstrate significant gains in communication competence, per self-ratings and clinician-ratings, upon completion of a communication-centered treatment, or CCT. The purpose of the present study was to determine whether gains in communication competence would also be reported by untrained observers. Eighty-one untrained observers completed an online survey that required each to view one of two videos depicting an adult who stutters during a mock interview recorded prior to CCT or after CCT. Participants were then asked to rate the communication competence of the interviewee on a 100-point visual analog scale and provide additional demographic information. Communication competence of the adult who stutters was rated significantly higher in their post-treatment video. Two observer-based factors were significantly associated with ratings of communication competence: years of education and years the respondent had known an adult who stutters. Upon controlling for these demographic factors, significantly higher ratings of communication competence for the post-treatment video were maintained. Although preliminary, findings suggest gains in communication competence demonstrated in previous studies through clinician and client observations are not limited to the sterile clinical environment, and further emphasizes the ecological validity of CCT. [*ClinicalTrials.gov* NCT05908123; https://clinicaltrials.gov/show/NCT05908123]

## Introduction

Public perception that adults who stutter are poor communicators is pervasive. Decades of research illustrate the widespread belief that effective communication – a skill that is considered essential for academic success (e.g., [1]), workplace advancement (e.g., [2–5]), and interpersonal relationships (see [6–7]) – cannot be adequately attained in the presence of stuttered speech. Based on this assumption, treatment options for adults who stutter have historically focused, in part or whole, on learning to speak fluently, and concealing moments of overtly stuttered speech (see systematic review by Brignell et al. [8]). The assumption that communication skills cannot be gained or improved without suppressing stuttered speech has gone unchallenged within clinical trials for decades. Only recently have clinical researchers demonstrated that targeting fluency during treatment is not necessary to improve the communication competency of persons who stutter.

Byrd and colleagues [9–11], for example, explored the impact of participation in a *communication-centered treatment (CCT)* designed to improve communication skills with no attempt to change speech fluency. Participants were rated to be significantly stronger communicators post-treatment by clinicians who were unfamiliar with the participants and blinded to pre-/post-treatment status of video samples [9–10]. Participants themselves also reported significantly stronger communication competencies after treatment across a variety of speaking contexts (dyad, small group, large group, public presentation) and listeners (strangers, acquaintances, friends; Coalson et al. [11]). Critically, neither self-nor clinician ratings of communication competency were predicted by pre-treatment stuttering frequency.

Taken together, these clinical data provide preliminary but compelling evidence that fluency and communication are not inextricably linked, at least from the perspective of speakers who stutter and their clinicians. Although promising, both the participants and the clinicians in the prior studies had shared knowledge of the nature of stuttering, the focus of CCT, and the desired clinical outcomes; thus, there was a relatively stable internal criteria for subjective evaluation, that one could argue may diverge from the appraisal of unfamiliar, untrained laypersons. Potential bias also may result from the sampling context. Clinicians who participated in the previous studies were blind to the pre-versus post-treatment status of videos, and videos were randomized, but they were evaluating a large number of consecutive videos depicting participants who stutter – a scenario rarely encountered in everyday life which may have also potentially compromised their ratings.

Therefore, to extend previous findings to a more ecologically valid context, the present study examined whether post-treatment gains in communication competency observed by clinicians in previous studies to reflect behavioral changes that are also identifiable to untrained, naïve observers. To do so, we recruited a large cohort of untrained observers to rate the communication competency of an unfamiliar adult who stutters based on a video sample recorded either before treatment had begun (Pre-treatment Video Stimuli) or after treatment had been completed (Post-treatment Video Stimuli). To explore implicit factors known or suspected to influence social evaluation, and evaluation of people who stutter (e.g., [12–13]) in particular, we also considered to what extent demographic and observer-related factors may account for perceived communication competency ratings for each video sample.

### Communication-Centered Treatment (CCT) for stuttering

The majority of treatment approaches for adults who stutter primarily or exclusively targeted fluency-centered speech techniques intended to either eliminate or minimize moment of stuttered speech (i.e., fluency shaping [14–15]; stuttering modification [16]). Yet, from the few randomized control trials (RCTs) that exist for adults who stutter, it is evident that fluency-centered treatment (a) has minimal impact on the psychological consequences of stuttering [17–18]), (b) is prone to high rates of relapse (71% [19]), and (c) may compromise the speaker’s innate ability to communicate (e.g., unnatural, effortful, and/or incongruent with their identity [20]). Additionally, listeners often rate speech techniques often employed during treatment to achieve fluency as equally or less desirable than stuttered speech [21–23].

Furthermore, several recent studies indicate that stuttering severity does not predict communication attitudes in person who stutter regardless of age (e.g., children [24]; adults: [25]). These data challenge the assumption that fluency must be targeted to facilitate positive perspectives of self and/or communication in persons who stutter. In fact, Byrd and colleagues [9-11, 26-27] provide evidence that significant positive changes in communication attitudes, and communication competence, can be reliably attained through participation in a treatment that focuses on improving overall communication and *explicitly* excludes clinical goals that attempt to hide, eliminate, or modify stuttered speech in children and adults following CCT.

### Preliminary findings with children who stutter

Byrd et al. [26, n = 23, ages 7- to 14-years old] examined changes in cognitive and affective wellbeing before and after treatment reported by 23 children and adolescents who stutter and their parents. Specifically, adolescents who stutter reported greater quality of life (as measured by the Overall Assessment of Speaker’s Experience with Stuttering [28]) following treatment, and parents reported significant improvement in their child’s ability to establish peer relationships (as measured by the PROMIS-Pediatric Short Form Peer Relationships Scale [29]).

A follow-up study by Byrd et al. [27] replicated these findings in an additional 23 child and adolescent participants (ages 7- to 14-years old). That is, participants and their parents reported significant post-treatment gains in quality of life and peer relationships. Combined, these findings indicate that treatment that excludes any attempt to modify speech fluency of children who stutter, and instead targets communication skills, results in significant gains that meet or exceed those previously reported for fluency-focused or stuttering modification treatment approaches.

Byrd et al. [9] extended analyses of their communication centered, whole person approach, by examining communication competencies in 37 children and adolescents who stutter (ages 4- to 17-years old) pre-versus post-treatment. An unfamiliar clinician rated pre- and post-treatment presentations (3 to 4 minutes in length), recorded in front of a large group of peers, based on nine difference communication skills: (1) language use, (2) language organization, (3) speech rate, (4) intonation, (5) volume, (6) gestures, (7) body position, (8) eye contact, and (9) facial affect (for detailed description, see Byrd et al. [9–10]). Findings provided preliminary evidence that, in addition to replicating the positive post-treatment changes in cognitive and affective aspects of stuttering reported in prior studies (Byrd et al. [26–27]), clinicians rated communication competency of children and adolescents who stutter during presentations as significantly stronger in samples recorded after treatment. Of particular relevance to the present study, these changes in communication competence following CCT were not significantly predicted by pre-treatment stuttering frequency.

### Preliminary findings with adults who stutter

Positive post-treatment gains in communication competence after treatment reported for children who stutter have been replicated in adults who stutter who have participated in CCT. Coalson et al. [11] examined self-reported clinical outcomes from 33 adults who stutter after an 11-week communication-centered treatment (for greater detail see Byrd et al. [10]) program similar to the one-week treatment program for children described in Byrd et al. [9, 26–27]). During the first and last week of treatment, participants completed the Self-Perceived Communication Competence [30] - a brief scale designed to self-assess communication skills in four specific communicative contexts (dyad, small group, large meeting, presentation) with three interlocutors (stranger, friend, acquaintance). Significant gains in self-rated communication competence were reported post-treatment across all 12 speaking scenarios and, similar to the children and adolescents who stutter in Byrd et al. [9], post-treatment gains were not predicted by stuttering frequency.

Improvement in communication competence is not limited to the perspective of the speaker who stutters. Byrd et al. [10] examined post-treatment communication competencies in 11 adults who stutter who participating in communication-centered treatment. Each participant completed a mock interview with an unfamiliar interviewer during the first week (pre-treatment sample) and final week (post-treatment sample). Randomized video samples of these interviews were rated offline by an unfamiliar speech-language pathologist blind to pre-/post-treatment status of each video. As observed for children and adolescents who have participated in CCT, adults who stutter demonstrated observable post-treatment improvements in eight of the nine targeted communication competencies (i.e., language use, language organization, speech rate, intonation, volume, gestures, body position, eye contact, and facial affect), and, again, these improvements were not predicted by pre-treatment stuttering frequency. Taken together with Coalson et al. [11], these preliminary data suggest that expert clinicians, as well as the adults who stutter themselves, observe positive changes in communication competencies when completing CCT, irrespective of pre- and/or post-treatment stuttering severity.

### Untrained observers

Although the client- and clinician-based outcome measures used in Byrd et al. [9–11] are commonplace within clinical trials of adult stuttering treatment (e.g., [17, 31–34]; however, see [35–38] for third-party ratings of naturalness), it could be argued that the changes reported were evaluated from two parties – the client and the clinician – whose shared perspectives invite a potential for rater bias because of their personal history and knowledge of stuttering and/or stuttering treatment. A logical means to address potential rater biases due to familiarity with the condition, and/or its treatment, is to examine clinical outcomes from the perspective of raters who have neither – the naïve observer.

To date, no clinical trials known to the present authors have investigated post-treatment *communication competence* for adults who stutter from the perspective of the naïve observer. Unlike clinicians or participants, untrained observers provide a valuable means to assess the ecological validity of any communicative outcome measure, by virtue of their inherently variable standards of communication competence. Novice laypersons rely upon a range of intrinsic and extrinsic cues to evaluate the quality of a speaker’s communication competence that are likely dissimilar from speakers and well-trained clinicians. By assessing the perspective of a large group of untrained observers, we can capture the variance of such internal criteria while also measuring the ecological impact of CCT outcomes. Thus, our primary research question is to assess to what extent the gains in communicative competence observed by clinicians in previous studies are also evident to general public. That is, we will analyze whether treatment outcomes observed by the trained clinician withstand the inherent variability of observer judgement.

### Observer-based variables

Naïve observers within the general public hold a well-documented negative bias towards persons who stutter (e.g., [39–42]). A number of demographic factors have been found to influence an observer’s evaluation of any speaker (e.g., age, gender, education, occupation, familiarity with language/multilingualism), including those who stutter (see [13, 43–44]). Such demographic factors as well as additional observer-based factors have a potential or documented influence on naïve observers’ attitudes towards adults who stutter (e.g., familiarity with person who stutters [45–46]; personal history with a communication disorder [47–48]; visible and/or nonvisible disability [49]).

Additionally, factors known to mitigate an observer’s overall evaluation of an adult who stutters as a *person* may override any attempt to measure a targeted trait, such as communication abilities, of specific communicative traits, resulting overly positive evaluations (see Werle & Byrd [50] for positive feedback bias by professors when evaluating presentations students who stutter) or overly negative evaluations (see Byrd et al. [12], for gender bias towards adults who disclosure stuttering). Given the central nature of communication abilities to how a person who stutters is perceived to the general public, it is worthwhile to examine whether ratings of communication competence – the focus of the present study – are attributable to public biases. Said another way, it is possible that observer ratings of the communication competence of a particular adult who stutters may be driven entirely by their overall perception of all people who stutter irrespective of their communicative skills. Thus, a second critical question to consider is to what extent observer-based factors mediate the opinions of the general public when rating communication abilities in adults who stutter to discern the unique influence of communication competence from generalized biases – positive or negative – towards adults who stutter.

### Rationale for the present study

The purpose of the present study was to examine how naïve observers rate the communication skills of an adult who stutters who completed an approach to treatment that focuses on communication effectiveness and makes no attempt to modify fluency or reduce stuttered speech. A secondary aim of this study was to assess whether observer-based factors of untrained observers influence perceived communication competence.

**RQ1:** Does communication effectiveness training yield positive gains in naïve observers’ evaluation of the communication skills of an adult who stutters?

**RQ2**: Do observer-based variables predict evaluation of the communication skills of an adult who stutters?

## Methods

The following study was approved by the authors’ university institutional review board (IRB: 2015-05-0044) and is part of an ongoing series of registered clinical trials (clinicaltrials.gov, NCT 05908123 [51]) designed to examine clinical outcomes of the Blank Center CARE Model^™^. Consent was obtained by all survey respondents prior to viewing communication competence stimuli. Communication competency stimuli was comprised of two separate videos: one depicting a speaker before he had completed CCT (Pre-treatment Video Stimuli) and one depicting the same speaker after he had received CCT (Post-treatment Video Stimuli).

### Communication-Centered Treatment (CCT)

A detailed description of the treatment program is provided in Byrd et al. [10]. The over-arching goal is to ensure individuals who stutter communicate effectively, advocate for themselves in a manner that maintains agency, and ensure their quality of life does not depend on producing, or attempting to control, stuttered speech. In brief, adult participants complete 11 weeks of treatment comprised of two 60-minute sessions per week (one group session, one individual session), totaling 22 sessions which include training in Communication, Advocacy, Resilience, and Education (the Blank Center CARE Model^™^). With respect to Communication, participants receive explicit instruction on how to appropriate incorporate nine core communication skills that do not rely on fluency speech production (i.e., language use, language function, speech rate, intonation, volume, gestures, body position, eye contact, facial affect). Training provided during the individual sessions provide a natural foundation for the weekly group sessions, wherein participants apply these skills in a variety of functional yet challenging speaking scenarios, including mock job interviews, small group mingling, impromptu icebreakers, one-on-one interactions with unfamiliar persons, and multiple presentations varied both in purpose (e.g., informative, persuasive, inspirational) and audience composition (e.g., small and large groups, familiar and unfamiliar listeners).

### Communication competency stimuli

Two video samples were selected from the 22 unscripted, impromptu mock interviews generated before and after treatment examined by Byrd et al. [10] – one recorded pre-treatment and its post-treatment counterpart. Each of the two video stimuli depicted a one-one-one, in-person mock interview between (a) an adult who stutters, who served as the interviewee, and (b) an unfamiliar clinical staff member of the Blank Center, who served as the interviewer. The same adult who stutters served as interviewee in both Pre-treatment and Post-treatment Video Stimuli. Interviewers differed between video stimuli to limit potential habituation to speaker. Each interviewer was unfamiliar with the participant who served as the interviewee, and both were provided identical, commonplace interview questions as prompts (e.g., “What do you consider your strengths and weaknesses?”; “Describe a prior work-related issue and how you addressed it.”).

Selection criteria included (a) significant intra-speaker gains in communication competence as rated by the speech-language pathologist evaluator, and (b) relatively comparable stuttering frequency and severity. Although selecting a pre- and post-video sample of the same participant with identical stuttering frequency and severity was not possible, as unscripted interactions naturally vary in length, stuttering frequency, and stuttering severity, the video samples selected for the present study were matched as closely as possible. As detailed in Table 1, stuttering severity equivalents for both the Pre- and Post-treatment Video Stimuli were rated as *moderate* per the Stuttering Severity Index – 4^th^ Edition (SSI-4 [52]) Stuttering frequency and severity ratings for each video were completed by an unfamiliar speech-language pathologist trained in disfluency count scoring but unfamiliar with the participant and the timepoint (Pre-treatment, Post-treatment) of each video. To further examine perceptual differences between video samples, a validation survey (administered as part of a separate project, Byrd et al. [53]) asked a separate cohort of naïve observers not included in this study to rate stuttering severity using a 100-point visual analog scale (0 = no stuttering, 100 = extremely severe stuttering) after rating one of the two video samples. Naïve observers rated stuttering severity to be statistically comparable (*p* = .95) between Pre- and Post-treatment Video Stimuli, with nearly identical mean severity ratings (*M* = 64.71 and 64.95, respectively; see Table 1).

**Table 1.** Characteristics of Pre-Treatment and Post-Treatment Video Stimuli.

|  | Pre-treatment Video<br>Stimuli | Post-treatment Video<br>Stimuli |
| --- | --- | --- |
| %SS<sup>a</sup> | 10.60% | 8.24% |
| Total syllables | 594 | 1258 |
| Total words | 453 | 959 |
| SSI-4<sup>b</sup> | Moderate | Moderate |
| Frequency | 7 | 6 |
| Duration | 10 | 10 |
| Physical Concomitants | 10 | 9 |
| Total | 27 | 25 |
| Observer-Rated Severity (100-point<br>VAS) |  |  |
| *M* (SE) | 64.71 (2.54) | 64.95 (2.41) |
| *N* | 63 | 57 |
| Length of Video | 5 min, 30 sec | 6 min, 25 sec |

<sup>a</sup>Percent of stuttered speech, disfluency types based on Yairi and Ambrose [54]
<sup>b</sup>Stuttering Severity Instrument-4<sup>th</sup> Edition [52]

The participant who served as the interviewee in the selected Pre- and Post-treatment videos was an adult Hispanic male who stutters. The participant spoke English and described himself as monolingual. No additional communication, developmental, psychological, neurological, and/or physical issues were reported by the participant or identified by the clinician during initial evaluation.

### Survey administration and respondent description

The two selected video samples were embedded in a Qualtrics-based survey distributed to adult, untrained observers via MTurk platform who were compensated for their participation, with the survey prompting one of the two videos in succession of access of the survey to ensure random observation of the Pre- and the Post-treatment sample. Each survey began with an informed consent landing page, followed by the instructions: “You are about to watch a video of an interview. Immediately following the video, you will be asked questions about the interviewee. The interview will be approximately 5 to 7 minutes in length. You will only be able to move forward in the survey after you have watched the video in its entirety.” Participants then watched either the video of the participant before training (Pre-treatment Video Stimuli), or the participant after training (Post-treatment Video Stimuli), with the “advance” button disabled for both the survey portal and the embedded video. Following the video, respondents were provided the following instructions accompanied by a 0-100 visual analog rating scale: “Using the scale below, please rate the interviewee’s communication skills. 0 = Communication skills not at all effective, 50 = Communication skills somewhat effective, 100 = Communication skills extremely effective.” Participants were then asked to describe what factors led to their rating in a free response text box. Following their rating and written feedback, respondents were asked to provide demographic information (e.g., age, race, ethnicity, gender, education, occupation, primary language). Participants were also asked to report their personal relationship with stuttering, persons who stutter, or other communication disorders (“Are you a person who stutters? Do you personally know a person who stutters? If so, please describe your relationship and how long you have known this person. Have you had previous speech, language, and/or hearing evaluation or therapy?”) as well as any visible or nonvisible diagnoses unrelated to communication difficulties (i.e., physical condition, psychological condition, neurological condition, emotional condition, vision/hearing loss, reading disorder, other/describe, none). Each survey included three attention check questions and four comprehension check questions to assess quality of individual responses. Survey data was collected in July 2021, and all respondents who completed the survey following the Pre-treatment and Post-treatment Video Stimuli in full were paid per MTurk standards of distribution.

The survey was initiated by 128 respondents (67 Pre-treatment Video Stimuli, 61 Post-treatment Video Stimuli). Of these 128 respondents, 36 (28%, 19 Pre-treatment Video Stimuli, 17 Post-treatment Video Stimuli) did not pass at least one of the seven attention/comprehension check questions during the course of the survey and were excluded from final analysis. Of the remaining 92, four were excluded because they self-identified as a person who stutters (2 Pre-treatment Video Stimuli, 2 Post-treatment Video Stimuli). Seven additional respondents were excluded (3 Pre-treatment Video Stimuli, 4 Post-treatment Video Stimuli) due to free-response items that suggested unclear understanding of the task (e.g., “*She [the interviewer] can ask some more questions*.”), questionable attention to the study (e.g., “*Everything is perfect*.”), or potentially auto-generated responses (e.g., unusual format of free-responses repeated across items or participants). The final corpus included 81 respondents (43 Pre-treatment Video Stimuli, 38 Post-treatment Video Stimuli). See Table 2 for detailed description of respondents.

**Table 2.**
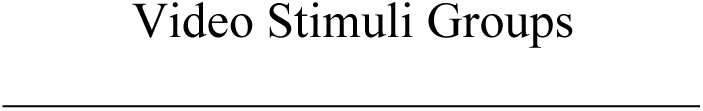
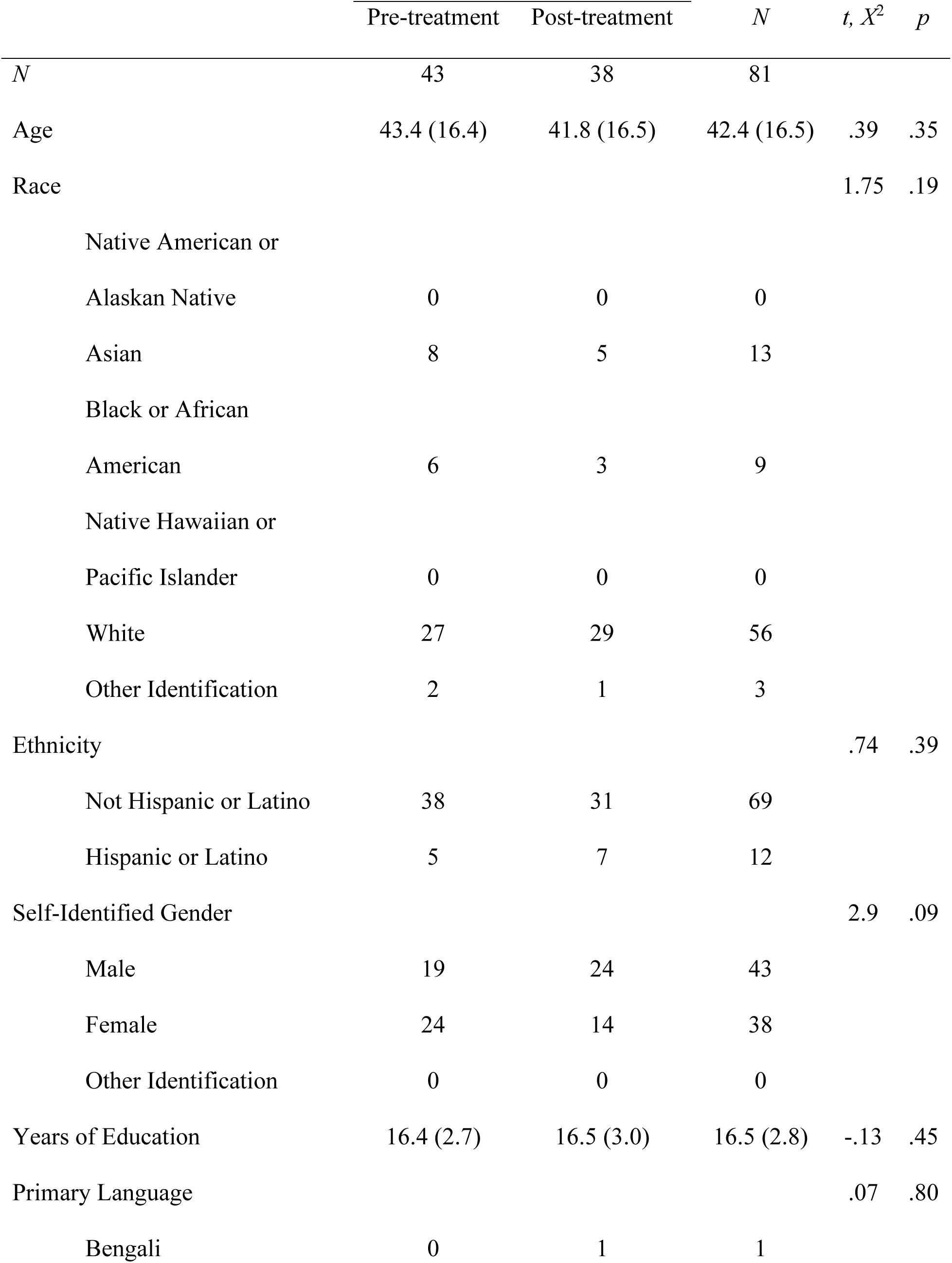
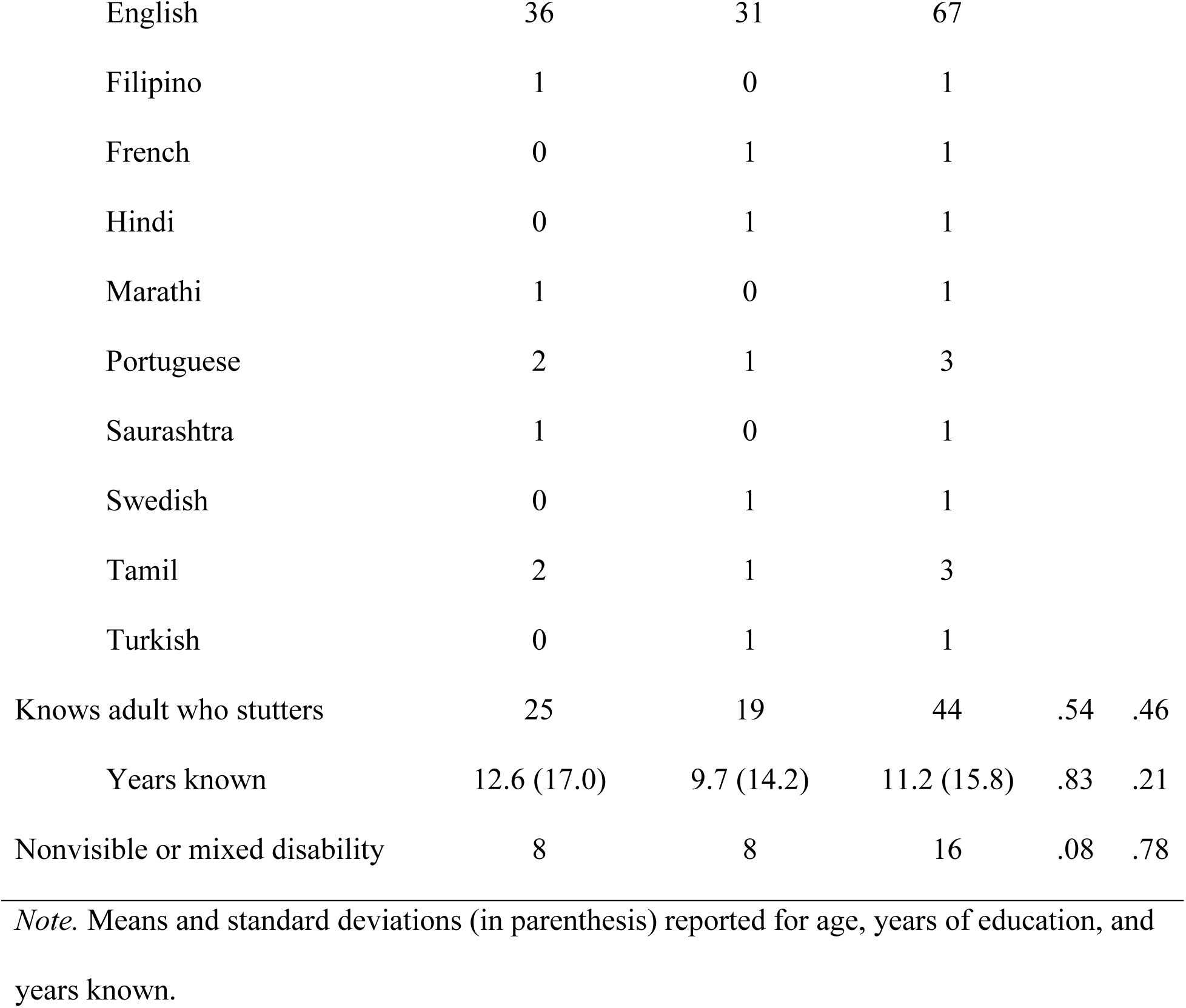
Demographic characteristics of naïve observer groups.

|  | Pre-treatment | Post-treatment | *N* | *t, X<sup>2</sup>* | *p* |
| --- | --- | --- | --- | --- | --- |
| *N* | 43 | 38 | 81 |  |  |
| Age | 43.4 (16.4) | 41.8 (16.5) | 42.4 (16.5) | .39 | .35 |
| Race |  |  |  | 1.75 | .19 |
| Native American or |  |  |  |  |  |
| Alaskan Native | 0 | 0 | 0 |  |  |
| Asian | 8 | 5 | 13 |  |  |
| Black or African |  |  |  |  |  |
| American | 6 | 3 | 9 |  |  |
| Native Hawaiian or |  |  |  |  |  |
| Pacific Islander | 0 | 0 | 0 |  |  |
| White | 27 | 29 | 56 |  |  |
| Other Identification | 2 | 1 | 3 |  |  |
| Ethnicity |  |  |  | .74 | .39 |
| Not Hispanic or Latino | 38 | 31 | 69 |  |  |
| Hispanic or Latino | 5 | 7 | 12 |  |  |
| Self-Identified Gender |  |  |  | 2.9 | .09 |
| Male | 19 | 24 | 43 |  |  |
| Female | 24 | 14 | 38 |  |  |
| Other Identification | 0 | 0 | 0 |  |  |
| Years of Education | 16.4 (2.7) | 16.5 (3.0) | 16.5 (2.8) | -.13 | .45 |
| Primary Language |  |  |  | .07 | .80 |
| Bengali | 0 | 1 | 1 |  |  |
| English | 36 | 31 | 67 |  |  |
| Filipino | 1 | 0 | 1 |  |  |
| French | 0 | 1 | 1 |  |  |
| Hindi | 0 | 1 | 1 |  |  |
| Marathi | 1 | 0 | 1 |  |  |
| Portuguese | 2 | 1 | 3 |  |  |
| Saurashtra | 1 | 0 | 1 |  |  |
| Swedish | 0 | 1 | 1 |  |  |
| Tamil | 2 | 1 | 3 |  |  |
| Turkish | 0 | 1 | 1 |  |  |
| Knows adult who stutters | 25 | 19 | 44 | .54 | .46 |
| Years known | 12.6 (17.0) | 9.7 (14.2) | 11.2 (15.8) | .83 | .21 |
| Nonvisible or mixed disability | 8 | 8 | 16 | .08 | .78 |

*Note.* Means and standard deviations (in parenthesis) reported for age, years of education, and years known.

### Analyses

#### RQ1

An independent *t*-test was conducted to compare naïve observers’ evaluation of communication competency depicted in one of two stimuli - a video depicting an adult who stutters who received communication effectiveness training (Post-treatment Video Stimuli) or a video depicting an adult who stutters who has not received communication effectiveness training (Pre-treatment Video Stimuli). Video type (Post-treatment Video Stimuli, Pre-treatment Video Stimuli) served as the independent variable, and ratings from the 100-point visual analog scale (VAS) of communication skills served as the dependent variable (0 = low competence, 100 = high competence). Effect sizes were obtained using Cohen’s *d* [55]. Because of the preliminary nature of the data and modest sample size, findings were verified by non-parametric analysis (Mann-Whitney *U*, α = .05).

#### RQ2

A linear regression was conducted to assess the influence of viewing communication competency stimuli (Post-treatment Video Stimuli, Pre-treatment Video Stimuli) and nine observer-related variables (age, race, ethnicity, gender, years of education, non-English primary language, knowing an adult who stutters, number of years respondent has known adult who stutters, nonvisible diagnosis; see Table 2) upon ratings of the communication skills of an adult who stutters. Categorical variables with responses that were either not reported (i.e., non-binary self-identified gender) or reported infrequently (i.e., non-English primary language with fewer than 4 respondents) were transformed to create a single binary variable (i.e., male/female; English/non-English primary language). To maintain relatively even distribution amongst categories during analysis, race was collapsed into a single binary variable (i.e., White, people of color) due to relatively infrequent self-identification as Black/African American (n = 9; Pre-treatment Video Stimuli = 6, Post-treatment Video Stimuli = 3) or racial identification that was not included in existing categories (n = 3, Pre-treatment Video Stimuli = 2, Post-treatment Video Stimuli = 1).

To determine which of the nine observer-related factors held meaningful predictive value of observer ratings, and therefore qualify for entry into the linear regression, we applied a version of Hosmer et al.’s [56] step-by-step method for *purposeful selection of covariates* modified for OLS linear regression. First, nine univariate analyses were conducted for each variable (chi-square tests for categorical variables, independent t-tests for continuous variables). Only variables with *p*-values greater than 0.25 were excluded. Second, a model with non-excluded variables was fitted, then each predictor re-assessed and deleted if significance exceeded *p* > .05. Third, the reduced model was compared to the original model using *F* values to ensure improved fit and to verify that change in beta coefficients between models did not exceed 20% (i.e., deleting-refitting-verifying cycle). Fourth, any variables that were excluded during the initial step were re-entered into the model, one at a time, but retained only if *p*-values were less than .05. Fifth, any interaction terms of interest between the remaining variables were entered into the model. Interaction terms were assessed using the deleting-refitting-verifying cycle used for main effects and retained only if statistically significant at *p* < .05 and if model fitness improved. Any main effects and interaction terms remaining after these steps were completed comprised the final model (see Table 3). Because of the preliminary nature of the data and modest sample size, bootstrap analysis was conducted to confirm initial findings (95% confidence intervals; 5000 samples).

**Table 3.** Summary of regression analyses of stimuli (Pre-treatment Video Stimuli, Post-treatment Video Stimuli) and observer-based factors predicting communication skills of an adult who stutters, as rated by naïve observers.

| | Variable | *B* | 95% CI | $\beta$ | *t* | *p* | *F* | *df* | *R*<sup>2</sup> |
| --- | --- | --- | --- | --- | --- | --- | --- | --- | --- |
| Model 1 | Intercept | -5.25 | -11.34, .78 |  | -1.68 | .089 | 6.03 | 1, 79 | .071 |
|  | **Pre-/Post-treatment Video Stimuli** | **11.24** | **2.38, 20.11** | **.27** | **2.46** | **.013** |  |  |  |
| Model 2 | Intercept | -5.68 | -11.25, -.11 |  | -1.95 | .046 | 7.30 | 3, 77 | .221 |
|  | **Pre-/Post-treatment Video Stimuli** | **12.12** | **3.97, 20.26** | **.29** | **2.84** | **.004** |  |  |  |
|  | Years of education | -2.47 | -3.92, -1.03 | -.33 | -3.28 | <.001 |  |  |  |
|  | Years AWS known | .23 | -.03, .49 | .17 | 1.67 | .087 |  |  |  |
| Bootstrapped | Intercept |  | -11.59, .07 |  |  | .063 |  |  |  |
|  | **Pre-/Post-treatment Video Stimuli** |  | **3.88, 20.72** |  |  | **.007** |  |  |  |
|  | Years of education |  | -3.99, -.85 |  |  | .003 |  |  |  |
|  | Years AWS known |  | .01, .46 |  |  | .049 |  |  |  |

*Note.* CI = confidence interval for unstandardized beta coefficients; AWS = adult who stutters

### Results

#### RQ1

An independent *t*-test was conducted to assess how naïve observers rate communication skills of an adult who stutters. As depicted in Fig 1, findings reveal significantly stronger perceived communication skills when viewing the video of a speaker post-CCT (Post-treatment Video Stimuli; *M* = 70.3, *SD* = 21.1) than when viewing a video of a speaker pre-CCT (Pre-treatment Video Stimuli; *M* = 59.0, *SD* = 20.1), *t*(79) = 2.46, *p* = .016, *d* = .55 [medium effect size]. Findings were confirmed via nonparametric analysis, *U*(43,38) = 532.50, *z* = 2.70, *p* = .007.

**Fig 1.**
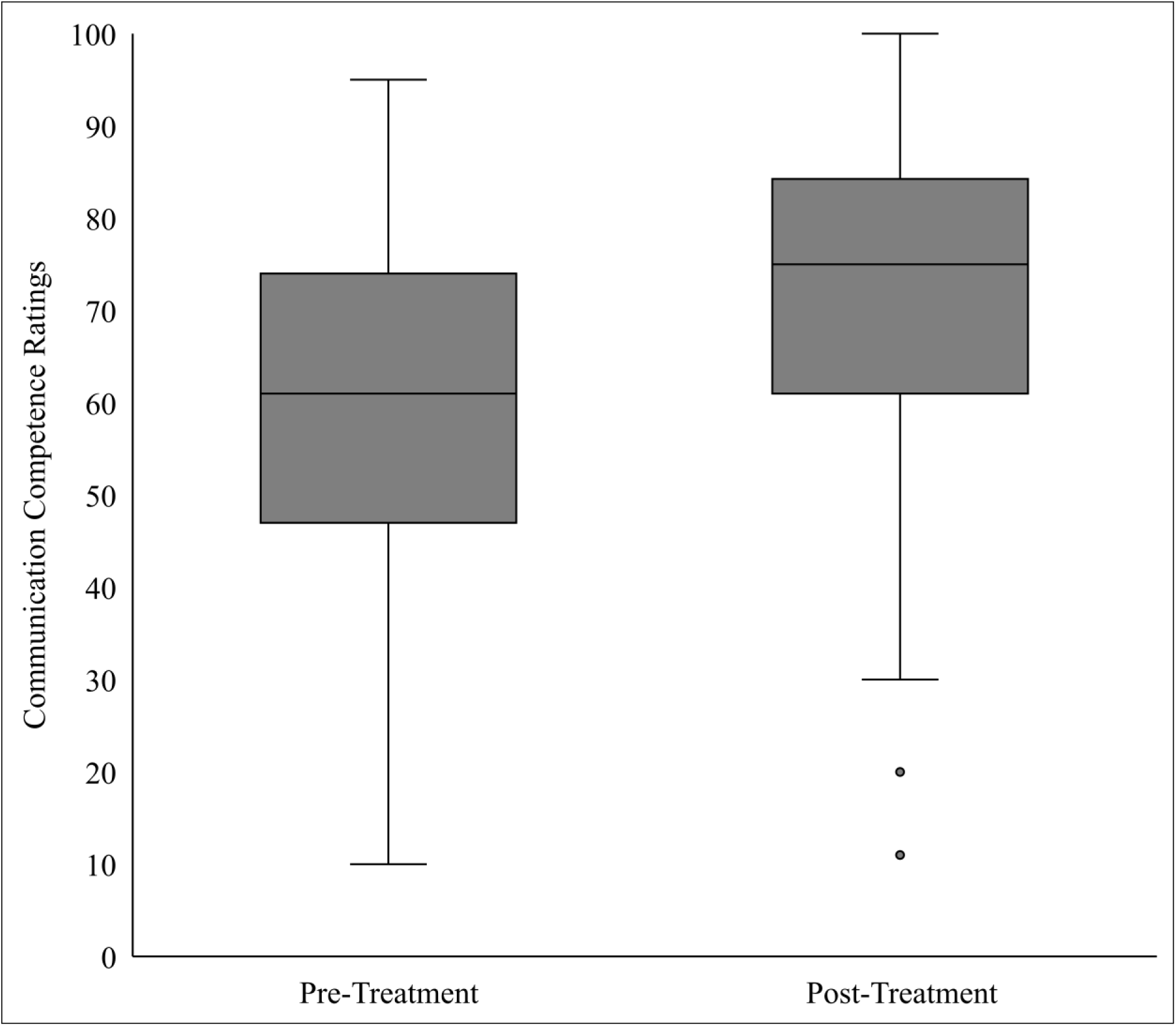
Communication competence of an adult who stutters, as rated by untrained observers before CCT (Pre-Treatment) and after CCT (Post-Treatment)

#### RQ2

A linear regression was conducted to determine the contribution of observer-related factors (i.e., age, race, ethnicity, gender, years of education, non-English primary language, knowing an adult who stutters, number of years respondent has known adult who stutters, nonvisible diagnosis) upon ratings of the communication skills of an adult who stutters. As expected, the video stimuli depicting a speaker post-treatment was a significant predictor of higher observer ratings when entered as the lone predictor variable (β = .67, *p* = .013), explaining 7.1% of the variance (*R*^2^ = .071; *F*(1, 79) = 6.03, *p* = .016; see Model 1 in Table 2). Upon completing Hosmer et al.’s (2013) purposeful selection of covariates, only two factors were identified as potential predictive covariates: (1) years of education, which significantly predicted observer ratings (β *= -*.33, *p* < .001) and accounted for 12.2% of the variance (*R*^2^ = .122), and (2) years the respondent has known an adult who stutters, which approached significance (β *= .*17*, p* = .087), and accounted for 2.8% of the variance (*R*^2^ = .028). After accounting for the contribution of these two observer-based factors, video stimuli of a post-treatment speaker remained a significant, positive predictor of improved observer ratings (β *=* .29*, p* = .004) with the final model accounting for 22.1% of the variance (*R*^2^ = .221; *F*(3, 77) = 7.30, *p* < .001; see Model 2 in Table 3).

To verify these outcomes, a bootstrapping analysis was conducted to determine 95% confidence interval (CI) for unstandardized beta coefficients of each factor based on 5000 samples. Bootstrap analysis confirmed a significant, positive coefficient for video stimuli depicting a post-treatment speaker (*p* = .007, [CI: 3.88, 20.72]) while controlling for potential influence of both observer-related factors (years of education: *p* = .003, [CI: -3.99, -.85]; years respondents have known an adult who stutters: *p* = .049 [CI: .01, .46]).

## Discussion

The primary purpose of this study was to examine whether naïve observers rated an adult who stutters as a stronger communicator after completing a specialized 11-week treatment program (CCT) designed to enhance communication skills rather than stuttering frequency or severity. A secondary purpose was to examine to what extent observer-related factors mediated these ratings. A large sample of naïve observers were recruited to view, and then rate, one of two videos of an adult who stutters who had completed the treatment program (Post-treatment Video Stimuli) or who had yet to complete the treatment program (Pre-treatment Video Stimuli). Findings indicate that naïve observers rated the speaker in the Post-treatment Video Stimuli to be a stronger communicator than the same speaker in the Pre-treatment Video Stimuli video. Two observer-rated factors were identified as significantly associated with communication competency ratings: (1) years of rater education and (2) years the rater had personally known an adult who stutters. Accounting for these factors, naïve observers nevertheless rated the video of the adult who stutters recorded after training as a significantly stronger communicator.

### RQ1: Communication competence gains from the perspective of naïve observers

Naïve observers rated the communication competence of an adult who stutters demonstrates during a mock interview recorded after CCT (Post-treatment Video Stimuli) as significantly higher than the video of the same adult, in the same mock interview setting, recorded prior to CCT (Pre-treatment Video Stimuli). These findings are consistent with the significant pre-/post-treatment gains in communication competence reported for adults who stutter as rated by speech-language pathologists [10] and client’s self-reported dyadic interactions ([11]; dyadic interactions: *p* < .0001). Findings also corroborate significant gains in communication competence observed by speech-language pathologists for young children and adolescents after a one-week treatment program based on the same clinical principles [9]. Consistency across ratings of communication competence from three difference perspectives – client, clinician, and naïve observer – provide confidence that changes in previous studies were not attributable to rater bias (i.e., client or clinician) and perhaps reflect a meaningful change in communication abilities also observable by untrained laypersons.

Unlike previous studies by Byrd and colleagues [9–10], wherein a single clinician rated pre-/post-treatment video samples from multiple adults who stutter (i.e., many-to-one), the structure of observation in this study was reversed. In those previous studies, our analyses captured the variance of treatment outcomes across multiple participants, with rater variance held constant by use of a single clinician rater. In the present study, multiple observers rated communication competence of a single adult who stutters (i.e., one-to-many) based on viewing one of two videos. By assigning one video to one respondent per survey, we were able to minimize respondent burden of watching multiple videos and potential order effects. Moreover, this statistical design allowed us to capture the variance of responses amongst the general public in response to changes in communication competence previously observed by clinicians. These data provide preliminary proof that gains observed in the clinic may also generalize to the perceptions of unfamiliar observers beyond the clinic – an issue of critical concern for any clinical trial and many participants in treatment. That being said, it is possible that the consistency between within-clinic and beyond-clinic evaluation of communication competence observed in this study may vary between individual participants. A natural next step is to assess the variance of treatment outcomes across participants by a single naïve observer, similar to one-to-many rating design in Byrd et al. [9–10], practical issues notwithstanding (e.g., order effects, participant/respondent burden). For now, findings from the present study provide meaningful external validity for the positive outcomes a novel treatment approach, and suggest that gains in communication competence, when observed in the clinic by a single clinician, will likely be observed by the general public.

As noted, stuttering frequency and severity in the Pre-Treatment and Post-treatment Video Stimuli were comparable (Pre-treatment Video Stimuli: SSI-4: 31 moderate [score 27]; Post-treatment Video Stimuli: SSI-4: moderate [score 25]), lending support to the notion that ratings of communication competence were not driven by changes on overt or observable stuttering behaviors. One may suggest that, similar to the rationale of the present study for communication competence, ratings of stuttering severity provided by expert clinicians may not reflect judgments of severity by the naïve listeners in the general public, and potentially influenced the observed-based ratings of communication competence more than expected. Comparable observer-based ratings of stuttering severity for Pre-Treatment and Post-treatment Videos reported in Table 1 (*p* = .95) suggest this was not the case. It is also possible that observer judgment of stuttering severity was less distracting because it was accompanied by higher (or lower) communication abilities. For example, Werle et al. [57] found that naïve observers who view a presenter who producing 15% SLDs but demonstrates high communication competence rate the stuttering as less distracting than the same presenter produced 15% SLDs but demonstrated low communication competence.

Further assessment of qualitative feedback from respondents in the present study support the findings of Werle et al. [57] and indicate that fluency was less of a concern in the presence of stronger communication competence in the Post-treatment Video Stimuli. When comparing the most neutral observers (i.e., 50% who provided the neither the most or least favorable ratings in each group; 25^th^ to 75^th^ percentile), respondents who watched the Post-treatment Video Stimuli noted that they certainly heard the interviewee stuttering, but also commented that its importance was offset by communication skills (e.g., “*The interviewee was very articulate and concise in his language and tone of voice. He used his hands when speaking, which made him appear more animated and that was easier to follow.”* “*Even though the interviewee had a stuttering issue, he was able to explain himself well. He gave good examples when asked for them by the interviewer*.”; “*I believe that [he] communicated well. … He looked the interviewer directly in the eyes, smiled, and nodded*.”). Accordingly, respondents who viewed the Pre-treatment Video Stimuli (25^th^ to 75^th^ percentile) often focused either on stuttering alone (e.g., “*He was not bold and confident about the way he deliver[s] things. He is a stammer[er]*”; “*The interviewee has a speech impediment and it is difficult for him to communicate verbally*.”) or stuttering in addition to poor nonverbal communication skills (e.g.. “*He seems to stay within his capabilities of communication, but his stutter is distracting. He answers questions directly, but he doesn’t use much eye contact.”*; *“He had a stutter and his body language looked tense but he gave great answers.*”) In sum, although similarly moderate stuttering was present in both videos and perceived by both respondent cohorts, qualitative data from the present study indicate that, consistent with Werle et al. [57], heightened communication skills of the speaker who received training helped to minimize the relevance of stuttering severity during observers judgment.

### RQ2: Observer-based factors associated with ratings of communication competence

Two demographic factors – years of education and years the respondent had known an adult who stutters - were identified as significant predictors of observer-rated communication competence. Specifically, observers rated communication competence to be stronger as the number of years the respondent had personally known an adult who stutters increased, but weaker as the amount of education the respondent had completed increased. Although observer-rated evaluations of communication competence remained significant upon controlling for these factors during analyses, the potential implications of these two factors warrant discussion. In terms of the number of years respondents have known a person who stutters, previous research has indicated that people who have developed first-hand relationships with any stigmatized groups are likely to report improved overall judgments of persons within that group (e.g., [58–59]), including persons who stutter (e.g., [60–61]; cf. [46, 62]). It is noteworthy that simply knowing/not knowing a person who stutters was insufficient to impact ratings, suggesting that the quantity and perhaps quality of time spent with an individual who stutters is necessary to significantly change how one views the communication abilities of a person who stutters. Future studies examining observer evaluations of participants who stutter should continue to include this as a measured factor, based on its long-standing influence on the general public views individuals who stutter.

Across videos, there was a significant negative relationship between years of education and communication ratings, wherein participants with higher levels of education often provided lower ratings of the speaker’s communication abilities, regardless of the video presented. To be clear, the mean number of years of education for each respondent group was equivalent (Pre-treatment Video Stimuli; *M* = 16.4 years of education; Post-treatment Video Stimuli, *M* = 16.5 years of education, *p* = .45). Research investigating responses to historically stigmatized groups has documented several patterns of responses held by evaluators. The pattern of response in the present study is consistent with the status characteristic model (e.g., [63]). Within this model, evaluators hold lower expectations of a particular group, and group members must perform exceptionally compared to in-group peers to achieve equivalent ratings. This pattern of response is in contrast to the positive feedback bias found in the shifting standards model in which lower expectations of the stigmatized group result in unduly earned higher ratings [64]. Relevant to the present study, researchers have found that when evaluators are judging individuals for maximum competency or skills, they align more with the status characteristic model [65]. In other words, when tasked with examining for top performance, evaluators rate members of stigmatized groups more critically and assign lower ratings. One interpretation of the present results is related to the video context and protocol directions. Job interviews are arguably high-stakes experiences in which candidates are evaluated critically. It is possible that, when tasked with evaluating communication skills within this high-stakes context, individuals with more years of education were evaluating more sensitively and/or with higher expectations than individuals with fewer years of education. This could possibly be anchored in their own personal experiences as either an interviewee or interviewer.

Results are contrary to recent works by Werle and Byrd [50, 66], however, who detailed a potential positive response bias when raters with higher years of education (i.e., college professors) evaluate students who stutter. It is possible that positive response bias becomes less evident for respondents who experience fewer years of academic evaluation based on communication abilities, or perhaps a unique pattern observed for professors and teachers whose job duties require ongoing evaluation of adult students. It is also possible that dyadic speaking context lowered the likelihood of positive feedback bias found for professors, who were perhaps more likely to overcorrect their personal biases observed for Werle and Byrd [50, 66] when presented with a speaker in a context for which they regularly provide evaluation (e.g., presentations). In that respect, positive response bias for the mock interviews included in the present study may be more apparent for respondents with a history of employment or training in human resources. Combined, although these two factors held significantly influence on ratings of communication competence, the significance of video status (Pre-treatment versus Post-treatment) beyond the influence of these factors, as well as the large number of demographic information included in the analyses, provides greater confidence that CCT training may be effective irrespective of unique demographic factor.

### Limitations and future studies

Although these combined factors accounted for an estimated 22.1% of the variance in the sample population of untrained observers – a non-trivial amount of inter-subject variability when evaluating communication competence of an adult who stutters - we acknowledge that a number of additional factors, known and unknown, beyond the focus of CCT also influence observer judgments. For example, visual nonverbal information (e.g., attire, physical appearance, environments) have a documented effect on evaluator judgments (e.g., [67]). To this point, it should be noted that the same adult was depicted in both video samples in the present study and, by coincidence, the participant happened to be wearing a necktie on during the first interview (Pre-treatment Video Stimuli). Although this may be considered more professional attire, this video was also rated less favorably, suggesting that attire was less critical than overall communication skills. Another unfortunate potential speaker-based factor that may influence ratings, but unrelated to CCT, was degree of accent and language proficiency. To be clear, the interviewee in this study was monolingual with no observable accent, nor did respondents note the speaker’s accent or English proficiency during subjective feedback. During pilot studies, however, naïve observers rated a multilingual adult who stutters with strong English proficiency but a heavier accent less favorably during a similar interview sample. These same respondents often provided specific negative feedback focusing on the speaker’s accent and/or level of English proficiency. Future studies of this nature should provide a clear definition of as well as linguistic versus speech fluency from the outset, similar to the instructions provided to respondents in Werle and Byrd [50, 66] fluency versus communication skills.

From a methodological standpoint, video samples that naturally vary in length and content also introduce potential confounds. For example, in the present study, the Post-treatment Video was longer and contained more total words than the Pre-treatment Video Sample, introducing the possibility that respondents became fatigued or impatient when viewing and prior to VAS rating. Again, the Post-treatment Video Sample received a significantly stronger mean rating than the Pre-treatment Video Sample, which tempers this concern. It is also possible that, although both mock interviews were unprompted events, and interviewers were unknown to the participants, the participant benefitted from the previous mock interview experience and felt at greater ease in the identical space provided for both interviews. Additional review of communication skills in a different space, or perhaps investigation of content overlap between interviews, would address this point. Yet another consideration is that dyadic interviews lend themselves to a specific, one-on-one style of interaction that favors certain adults who stutter more than others, and post-CCT gains may not necessarily generalize to other contexts such as presentations (however, see Coalson et al. [11] for post-CCT gains across all speaking contexts as rated by adult clients who stutter). It is important to note this is one entry in a series of clinical trials focused on the outcomes of CCT from a variety of perspectives (e.g., self, clinician, observer). A larger cohort of respondents is always necessary to corroborate preliminary results, and examination of clinical outcomes will continue across multiple contexts, from multiple perspectives, and with multiple measures, in future studies.

## Conclusion

This study examined the clinical outcomes of CCT – a specialized treatment for adults who stutter that focuses on communication rather than fluency – from the perspective of naïve observers. Results found that naïve observers rated a participant depicted in a video sample recorded after treatment as a significantly stronger communicator than a video of the same participant recorded prior to treatment. Findings provide corroborating evidence that clinical gains in communication competence rated by clinicians, and self-rated by participants, in previous studies was also observed for unfamiliar, untrained observers.

## Data Availability

All data produced in the present study are available upon reasonable request to the authors

## Acknowledgements

This project was supported by the foundational grant support funded to the Arthur M. Blank Center for Stuttering Education and Research and endowed support provided through the Michael and Tami Lang Stuttering Institute, the Dr. Jennifer and Emanuel Bodner Developmental Stuttering Laboratory, and the Dealey Family Foundation Stuttering Clinic awarded to the second author. The authors would like to thank Michael Mahometa for his assistance with statistical analysis and all survey participants.

## References

1. Morreale SP, Osborn MM, Pearson JC. Why communication is important: A rationale for the centrality of the study of communication. J Assoc for Commun Adm. 2000;29(1):1.

2. AC Nielsen Research Services. Research on employer satisfaction with graduate skills. Interim Report Canberra, Australia. 1998

3. Mikkelson AC, York JA, Arritola J. Communication competence, leadership behaviors, and employee outcomes in supervisor-employee relationships. Bus Prof Commun Q. 2015 Sep;78(3):336–54.

4. Payne HJ. Reconceptualizing social skills in organizations: Exploring the relationship between communication competence, job performance, and supervisory roles. J Leadersh Organ Stud. 2005 Jan;11(2):63–77.

5. Scudder JN, Guinan PJ. Communication competencies as discriminators of superiors’ ratings of employee performance. J of Bus Commun (1973). 1989 Jun;26(3):217–29.

6. Beebe SA, Beebe SJ, Redmond M, Geerinck T, Salem-Wiseman L. Interpersonal communication: Relating to others. 6th Can. Ed. Toronto: Pearson Canada; c2015.

7. Spitzberg BH, Cupach WR. Interpersonal skills. In: Knapp ML & Daly JA, editors. The Sage handbook of interpersonal communication. 4th ed. Los Angeles (CA); SAGE; 2011. p. 481–524.

8. Brignell A, Krahe M, Downes M, Kefalianos E, Reilly S, Morgan AT. A systematic review of interventions for adults who stutter. J Fluency Disord. 2020 Jun;64:105766. doi: 10.1016/j.jfludis.2020.105766. Epub 2020 Apr 30. PMID: 32438123.

9. Byrd CT, Winters KL, Young M, Werle D, Croft RL, Hampton E, Coalson G, White A, Gkalitsiou Z. The Communication benefits of participation in Camp Dream. Speak. Live.: An extension and replication. Semin Speech Lang. 2021 Mar;42(2):117–135. doi: 10.1055/s-0041-1723843. Epub 2021 Mar 16. PMID: 33725730.

10. Byrd CT, Coalson GA, Young MM. Targeting communication effectiveness in adults who stutter: A preliminary study. Top Lang Disord. 2022 Jan 1;42(1):76–93.

11. Coalson, GA, Byrd, CT, Werle D. Self-reported communication competence by adults who stutter following communication competence treatment (CCT). Poster session presented at: National Black Association for Speech, Language, and Hearing Conference 2023. Apr 20-22. Washington, DC.

12. Byrd CT, McGill M, Gkalitsiou Z, Cappellini C. The effects of self-disclosure on male and female perceptions of individuals who stutter. Am J Speech Lang Pathol. 2017 Feb 1;26(1):69–80. doi: 10.1044/2016_AJSLP-15-0164. PMID: 28056467.

13. Louis KO. Epidemiology of public attitudes toward stuttering. In Stuttering meets stereotype, stigma, and discrimination: An overview of attitude research 2015 (pp. 7–42). West Virginia University Press Morgantown, WV.

14. O’Brian S, Carey B, Onslow M, Packman A, Cream A. The Camperdown program for stuttering: treatment manual. Sydney: Australian Stuttering Research Center. 2010 Jan.

15. Kroll R. The Fluency Plus Program: an integration of fluency shaping and cognitive restructuring procedures for adolescents and adults who stutter. In: Guitar B & McCauley R, editors. Treatment of stuttering: Established and emerging interventions. Philadelphia(PA): Lippincott Williams & Wilkins; 2010. p. 277-311.

16. Van Riper C. The treatment of stuttering. Englewood Cliffs(NJ): Prentice Hall; 1973.

17. Menzies RG, O’Brian S, Onslow M, Packman A, St Clare T, Block S. An experimental clinical trial of a cognitive-behavior therapy package for chronic stuttering. J Speech Lang Hear Res. 2008 Dec;51(6):1451–64. doi: 10.1044/1092-4388(2008/07-0070). Epub 2008 Jul 29. PMID: 18664696.

18. Menzies R, O’Brian S, Packman A, Jones M, Helgadóttir FD, Onslow M. Supplementing stuttering treatment with online cognitive behavior therapy: An experimental trial. J Commun Disord. 2019 Jul-Aug;80:81-91. doi: 10.1016/j.jcomdis.2019.04.003. Epub 2019 May 2. PMID: 31100535.

19. Craig AR, Hancock K. Self-reported factors related to relapse following treatment for stuttering. Aust J Hum Commun Disord. 1995 Jun 1;23(1):48–60.

20. Cream A, Onslow M, Packman A, Llewellyn G. Protection from harm: the experience of adults after therapy with prolonged-speech. Int J Lang Commun Disord. 2003 Jan 1;38(4):379–95.

21. De Nardo T, Tetnowski JA, Coalson GA. Listener perceptions of stuttering and stuttering modification techniques. J Fluency Disord. 2023 Mar;75:105960. doi: 10.1016/j.jfludis.2023.105960. Epub 2023 Jan 27. PMID: 36736074.

22. Manning WH, Burlison AE, Thaxton D. Listener response to stuttering modification techniques. J Fluency Disord. 1999 Dec 1;24(4):267–80.

23. Von Tiling J. Listener perceptions of stuttering, prolonged speech, and verbal avoidance behaviors. J Commun Disord. 2011 Mar-Apr;44(2):161-72. doi: 10.1016/j.jcomdis.2010.09.002. Epub 2010 Sep 16. PMID: 20947094.

24. Winters KL, Byrd CT. Predictors of communication attitude in preschool-age children who stutter. J Commun Disord. 2021 May-Jun;91:106100. doi: 10.1016/j.jcomdis.2021.106100. Epub 2021 Apr 14. PMID: 33862497.

25. Werle D, Winters KL, Byrd CT. Preliminary study of self-perceived communication competence amongst adults who do and do not stutter. J Fluency Disord. 2021 Dec;70:105848. doi: 10.1016/j.jfludis.2021.105848. Epub 2021 Apr 8. PMID: 33895686.

26. Byrd C, Chmela K, Coleman C, Weidner M, Kelly E, Reichhardt R, Irani F. An introduction to camps for children who stutter: What they are and how they can help. Perspect ASHA Spec Interest Groups. 2016 Mar 31;1(4):55–69.

27. Byrd CT, Gkalitsiou Z, Werle D, Coalson GA. Exploring the effectiveness of an intensive treatment program for school-age children who stutter Camp Dream. Speak. Live.: A follow-up study. Semin Speech Lang. 2018 Nov;39(5):458–468. doi: 10.1055/s-0038-1670669. Epub 2018 Sep 19. PMID: 30231266.

28. Yaruss JS, Quesal RW. Overall assessment of the speaker’s experience with stuttering. Stuttering Therapy Resources, Inc; 2016

29. Dewalt DA, Thissen D, Stucky BD, Langer MM, Morgan Dewitt E, Irwin DE, Lai JS, Yeatts KB, Gross HE, Taylor O, Varni JW. PROMIS Pediatric Peer Relationships Scale: Development of a peer relationships item bank as part of social health measurement. Health Psychol. 2013 Oct;32(10):1093–103. doi: 10.1037/a0032670. Epub 2013 Jun 17. PMID: 23772887; PMCID: DPMC3865609.

30. Richmond VP, McCroskey JC. Communication: Apprehension, avoidance, and effectiveness. Pearson; 1997.

31. Cream A, O’Brian S, Jones M, Block S, Harrison E, Lincoln M, Hewat S, Packman A, Menzies R, Onslow M. Randomized controlled trial of video self-modeling following speech restructuring treatment for stuttering. J Speech Lang Hear Res. 2010 Aug;53(4):887–97. doi: 10.1044/1092-4388(2009/09-0080). Epub 2009 Dec 22. PMID: 20029053.

32. Erickson S, Block S, Menzies R, O’Brian S, Packman A, Onslow M. Standalone Internet speech restructuring treatment for adults who stutter: A phase I study. Int J Speech Lang Pathol. 2016 Aug;18(4):329–40. doi: 10.3109/17549507.2015.1101156. Epub 2015 Nov 7. PMID: 27063674.

33. Erickson S, Block S, Menzies R, Onslow M, O’brian S, Packman A. Standalone Internet speech restructuring treatment for adults who stutter: A pilot study. J Clin Pract Speech Lang Pathol. 2012;14(3):118–23.

34. Menzies RG, Packman A, Onslow M, O’Brian S, Jones M, Helgadóttir FD. In-Clinic and Standalone Internet Cognitive Behavior Therapy Treatment for Social Anxiety in Stuttering: A Randomized Trial of iGlebe. J Speech Lang Hear Res. 2019 Jun 19;62(6):1614–1624. doi: 10.1044/2019_JSLHR-S-18-0340. Epub 2019 May 20. PMID: 31112442.

35. Carey B, O’Brian S, Onslow M, Block S, Jones M, Packman A. Randomized controlled non-inferiority trial of a telehealth treatment for chronic stuttering: The Camperdown Program. Int J Lang Commun Disord. 2010 Jan-Feb;45(1):108–20. doi: 10.3109/13682820902763944. PMID: 19424889.

36. Carey B, O’Brian S, Onslow M, Packman A, Menzies R. Webcam delivery of the Camperdown Program for adolescents who stutter: A phase I trial. Lang Speech Hear Serv Sch. 2012 Jul;43(3):370–80. doi: 10.1044/0161-1461(2011/11-0010). Epub 2012 Jan 9. PMID: 22232423.

37. Carey B, O’Brian S, Lowe R, Onslow M. Webcam delivery of the Camperdown Program for adolescents who stutter: a phase II trial. Lang Speech Hear Serv Sch. 2014 Oct;45(4):314–24. doi: 10.1044/2014_LSHSS-13-0067. PMID: 25091362.

38. O’Brian S, Onslow M, Cream A, Packman A. The Camperdown Program: outcomes of a new prolonged-speech treatment model. J Speech Lang Hear Res. 2003 Aug;46(4):933–46. doi: 10.1044/1092-4388(2003/073). PMID: 12959471.

39. Guntupalli VK, Kalinowski J, Nanjundeswaran C, Saltuklaroglu T, Everhart DE. Psychophysiological responses of adults who do not stutter while listening to stuttering. Int J Psychophysiol. 2006 Oct;62(1):1–8. doi: 10.1016/j.ijpsycho.2005.11.001. Epub 2006 Jan 18. PMID: 16414137.

40. Hurst MI, Cooper EB. Employer attitudes toward stuttering. J Fluency Disord. 1983 Mar 1;8(1):1–2.

41. Ruscello DM, Lass NJ, Schmitt JF, Pannbacker MD. Special educators’ perceptions of stutterers. J Fluency Disord. 1994 Jun 1;19(2):125–32.

42. Van Borsel J, Brepoels M, De Coene J. Stuttering, attractiveness and romantic relationships: the perception of adolescents and young adults. J Fluency Disord. 2011 Mar;36(1):41–50. doi: 10.1016/j.jfludis.2011.01.002. Epub 2011 Jan 20. PMID: 21439422.

43. Chu SY, Unicomb R, Lee J, Cho KS, St Louis KO, Harrison E, McConnell G. Public attitudes toward stuttering in Malaysia. J Fluency Disord. 2022 Dec;74:105942. doi: 10.1016/j.jfludis.2022.105942. Epub 2022 Nov 7. PMID: 36395547.

44. Lefort MKR, Erickson S, Block S, Carey B, St Louis KO. Australian attitudes towards stuttering: A cross-sectional study. J Fluency Disord. 2021 Sep;69:105865. doi: 10.1016/j.jfludis.2021.105865. Epub 2021 Jul 22. PMID: 34380103.

45. Klassen TR. Perceptions of people who stutter: Re-assessing the negative stereotype. Percept Mot Skills. 2001 Apr;92(2):551–9. doi: 10.2466/pms.2001.92.2.551. PMID: 11361320.

46. White PA, Collins SR. Stereotype formation by inference: a possible explanation for the “stutterer” stereotype. J Speech Hear Res. 1984 Dec;27(4):567–70. doi: 10.1044/jshr.2704.567. PMID: 6521464.

47. Lass NJ, Ruscello DM, Pannbacker M, Schmitt JF, Middleton GF, Schweppenheiser K. The perceptions of stutterers by people who stutter. Folia Phoniatr Logop. 1995;47(5):247–51. doi: 10.1159/000266358. PMID: 8563776.

48. Zhang J, Kalinowski J, Saltuklaroglu T, Hudock D. Stuttered and fluent speakers’ heart rate and skin conductance in response to fluent and stuttered speech. Int J Lang Commun Disord. 2010 Nov-Dec;45(6):670–80. doi: 10.3109/13682820903391385. PMID: 20017588.

49. Zernitsky-Shurka E. Ingroup and outgroup evaluation by disabled individuals. J Soc Psychol. 1988 Aug;128(4):465–72. doi: 10.1080/00224545.1988.9713766. PMID: 2972885.

50. Werle D, Byrd CT. Professors’ perceptions and evaluations of students who do and do not stutter following oral presentations. Lang Speech Hear Serv Sch. 2022 Jan 5;53(1):133–149. doi: 10.1044/2021_LSHSS-21-00069. Epub 2021 Dec 3. PMID: 34861764.

51. Byrd, C. T. The Blank Center CARE Model™: A non-ableist approach to treatment. ClinicalTrials.gov identifier: NCT05908123. Updated June 18, 2023. https://www.clinicaltrials.gov/ct2/show/NCT05908123

52. Riley G. Stuttering severity instrument for children and adults. Pro-Ed; 1994.

53. Byrd CT, Werle D, Coalson GA. Observer-rated outcomes of Communication-Centered Treatment (CCT) for adults who stutter. Poster session presented at: Georgia Speech-Language-Hearing Association. 2023 Feb 24-25. Savannah, GA.

54. Yairi E, Ambrose NG. Early childhood stuttering for clinicians by clinicians: Pro-Ed; 2005.

55. Cohen J. Statistical power analysis for the behavioral sciences. Academic press; 2013 Sep 3.

56. Hosmer Jr DW, Lemeshow S, Sturdivant RX. Applied logistic regression. John Wiley & Sons; 2013 Apr 1.

57. Werle D, Byrd CT, Coalson GA. Impact of self-disclosure and communication competence on perceived listener distraction. J Commun Disord. 2023 Apr 25;103:106333. doi: 10.1016/j.jcomdis.2023.106333. Epub ahead of print. PMID: 37130470.

58. Allport GW. The nature of prejudice. Reading (MA): Addison-Wesley; 1958.

59. Pettigrew TF, Tropp LR. A meta-analytic test of intergroup contact theory. J Pers Soc Psychol. 2006 May;90(5):751–83. doi: 10.1037/0022-3514.90.5.751. PMID: 16737372.

60. Hughes CD, Gabel RM, Palasik ST. Examining the Relationship Between Perceptions of a Known Person Who Stutters and Attitudes Toward Stuttering. Can J Speech Lang Pathol Audio. 2017 Jul 1;41(3).

61. Hughes S, Gabel R, Irani F, Schlagheck A. University students’ perceptions of the life effects of stuttering. J Commun Disord. 2010 Jan-Feb;43(1):45–60. doi: 10.1016/j.jcomdis.2009.09.002. Epub 2009 Sep 23. PMID: 19836026.

62. Doody I, Kalinowski J, Armson J, Stuart A. Stereotypes of stutterers and nonstutterers in three rural communities in Newfoundland. J Fluency Disord. 1993 Dec 1;18(4):36.

63. Foschi M. Gender and double standards for competence. Gender, interaction, and inequality. 1992:181–207.

64. Biernat M, Manis M, Nelson TE. Stereotypes and standards of judgment. J Pers Soc Psychol. 1991 Apr;60(4):485.

65. Biernat M, Kobrynowicz D. Gender- and race-based standards of competence: lower minimum standards but higher ability standards for devalued groups. J Pers Soc Psychol. 1997 Mar;72(3):544–57. doi: 10.1037//0022-3514.72.3.544. PMID: 9120783.

66. Werle D, Byrd CT. College professors’ perceptions of students who stutter and the impact on comfort approaching professors. J Fluency Disord. 2021 Mar;67:105826. doi: 10.1016/j.jfludis.2020.105826. Epub 2020 Dec 25. PMID: 33360979.

67. Burgoon JK, Manusov V, Guerrero LK. Nonverbal communication. New York: Routledge; 2016 Jan 8.

